# Health literacy profiles correlate with participation in primary health care among patients with chronic diseases: A latent profile analysis

**DOI:** 10.1101/2025.03.31.25324983

**Authors:** Qi Xu, Baokai Wang, Li Zhang

## Abstract

**Objectives:** Given the persistent, complex, and diverse health needs of chronic disease patients, their health literacy and active participation in the process of primary health service delivery has gained increasing attention. This study aimed to identify the potential profiles of patient health literacy, examine the sociodemographic and health factors associated with these profiles, and investigate the relationship between health literacy profiles and patient participation in primary health care.

**Methods:** A cross-sectional survey was conducted in Shandong Province, China, from July 14 to August 20, 2023. 911 patients with chronic diseases were selected using a multistage stratified sampling method. The survey assessed sociodemographic and health characteristics, patient health literacy, and patient participation. We adopted latent profile analysis (LPA) to identify the categories of patient health literacy, and multiple linear regression was performed to assess the impact of health literacy profiles on patient participation.

**Results:** Three distinct latent profiles were identified: low health literacy group (Profile 1,10.98%), moderate health literacy group (Profile 2, 56.86%), and high health literacy group (Profile 3, 32.16%). Age, marital status, residency, occupational status, education level, personal monthly income, and health status had statistically significant effects on patient health literacy profiles (*P*<0.05). Patients with higher health literacy exhibited significantly greater participation in primary health care than those with lower health literacy (B = 3.544, p < 0.001).

**Conclusions:** This study’s findings highlight the heterogeneity of health literacy among patients with chronic diseases and underscore the need for tailored interventions to enhance patient health literacy. In addition, the findings offer valuable insights for enhancing health literacy in facilitating patient participation in primary health care.

## Introduction

Chronic diseases have emerged as the leading cause of mortality worldwide [1]. They are characterized by the prolonged duration and the need for long-term management, and the effectiveness of their prevention and control heavily relies on the accessibility and continuity of primary health care [2, 3]. International experience has consistently shown that primary health care institutions provide the most effective platform for implementing comprehensive chronic disease management [4–6]. Since 2009, China has significantly increased financial investment and implemented beneficial policies aimed to strengthen its primary health care system, which plays a central role in preventing and managing chronic diseases [7]. However, with the accelerating aging population and changing lifestyles in China, the incidence of chronic diseases has rising steadily, posing a major public health challenge that significantly affects residents’ health and quality of life [8–10]. Therefore, policymakers must develop and implement effective treatment strategies to enhance chronic disease management.

Patients with chronic diseases have persistent, complex, and diverse health service needs. Facilitating patient involvement in the process of health service delivery, and enabling physicians to understand individual’ requirements, preferences, and values and invite patients to co-design care planning are particularly valuable aspects to meet these needs [11, 12]. Furthermore, under the patient-centered service model, the patient-physician relationship has evolved into a collaborative partnership emphasizing mutual participation, where patient autonomy is respected, and both medical information and care decisions are jointly negotiated between health care providers and individuals [13, 14]. In particular, as chronic diseases become more prevalent, patient participation in health care process has emerged as a crucial determinant in achieving patient-centered care and improving disease management [15–17]. Consequently, the participation among patients with chronic diseases is receiving more attention than ever before.

Patient participation encompasses a broader range of activities, including sharing experiences, knowledge, and information, engaging in interpersonal interactions, mutual communication, and participating in decision-making and self-management [18]. Such participation often occurs during interactions with physicians, where patients can ask questions, express preferences and opinions in decision-making, and provide suggestions on their care [11]. When patients engage actively in health service delivery, their health-related needs, preferences, and recommendations are integrated into the service designs through effective communication with health care providers [13]. Patient participation has increasingly been recognized in great practical value at both the individual and organizational level by previous empirical studies. Encouraging patient participation helps to enhance patients’ knowledge about their symptoms and conditions [19], improve patients’ understanding of treatment options and potential outcomes [20], enrich patient experiences [15], and lead to better health outcomes [14] and quality of life [21]. In addition, the emphasis and facilitation of patient participation by organizations is associated with reduced decision-making conflict [22], and higher quality and safety of health service [23]. Considering the crucial role of patient participation, it is essential to investigate the determinants that shape patient participation in health care.

The potential factors influencing patient participation behaviors are complex and multifaceted. Effective participation in health care requires patients to accurately comprehend health-related information and follow instructions from health care professionals [22]. Patients with limited health literacy may struggle to express their expectations, beliefs, needs, and concerns for health care, seek health information, ask questions about medical issues, and fully understand treatment plans [24, 25]. Thus, it is clear that health literacy plays a vital role in patient participation. Health literacy refers to one’s ability to gather, process, understand, evaluate, and apply health-related information and services to manage their health and make appropriate health decisions [26]. Adequate health literacy enhances a person’s knowledge, motivation to communicate with health care professionals, and ability to comprehend, evaluate, and utilize health-related information effectively [27]. Patients with higher levels of health literacy show a greater tendency to understand health-related materials, engage in meaningful communication with health care providers, and actively participate in disease management [28, 29]. Extensive research has demonstrated that active participation in health care decision-making requires sufficient health literacy [30–33]. However, few studies have specifically explored the potential relationship between health literacy and patient participation in primary health care among individuals with chronic diseases.

Given the significance of patient participation in chronic disease management, it is particularly important to assess the extent of patient participation in the health care process, and identify ways to enhance the participation in health care among patients with chronic diseases. In addition, whether health literacy influences patient participation in primary health care remains unclear. Furthermore, previous studies measuring health literacy have predominantly relied on the aggregate or average scores, potentially resulting in large heterogeneity across diverse subgroups. Latent profile analysis (LPA) provides a person-centered analytical approach that describes the heterogeneity within various health literacy groups by categorizing individuals based on specific characteristics of different items. Therefore, the purpose of this study was to explore the latent profiles of health literacy using LPA, examine the impact of sociodemographic and health factors on these profiles, and investigate the relationship between health literacy profiles and patient participation. The findings will provide valuable insights into the great importance of health literacy among patients with chronic diseases and inform strategies for incorporating health literacy best practices to enhance patient participation in primary health care.

## Methods

### Study design and population

This cross-sectional survey was conducted from July 14 to August 20, 2023 in Shandong Province, a populous and economically developed region in eastern China. A multi-stage stratified sampling method was used to recruit participants across 16 prefectures in Shandong Province. In the first stage, two districts or counties were selected randomly from each prefecture. In the second stage, urban and rural areas were stratified, and one subdistrict (urban area) and one township (rural area) were selected randomly from each district or county. In the third stage, one community health service center (for urban areas) or one township hospital (for rural areas) was randomly selected from each chosen subdistrict or township, resulting in 64 primary health care institutions as research sites. In the final stage, investigators including graduate and undergraduate students from the School of Health Management at Binzhou Medical University conducted convenience sampling to survey residents within the service areas of the selected primary health care institutions. All surveys were administered by trained investigators who verbally presented each question of the questionnaire to the participants and recorded their responses.

The inclusion criteria were as follows: (1) aged 18 years or older; (2) ability to communicate effectively without language barriers; and (3) previously utilized health services at one of the selected institutions. The exclusion criteria were as follows: (1) inability to comprehend the questionnaire content; (2) presence of language barriers or cognitive impairments; and (3) no prior use of health services at the surveyed institutions. Participants who agreed to participate in the survey provided written consent after receiving an explanation of the study’s purpose and were assured of their right to withdraw at any time. All investigators were trained in professional interviewing skills to ensure quality control and data reliability. In addition, all collected data were anonymized and maintained with strict confidentiality.

The health literacy level among residents in Shandong Province in 2023 was 33.13% [34]. Based on the standard sample size estimation formula for the cross-sectional survey, the minimum sample size for our study was calculated to be 340, with a 95% confidence interval, an absolute error of 5%, and a refusal rate of 10%. We collected 3149 questionnaires, of which 128 were excluded due to logical errors or incomplete responses. Consequently, 3021 valid questionnaires were obtained, yielding a valid response rate of 95.94%. Based on self-reported responses to the question regarding chronic disease status, 911 participants (30.16% of the total sample) reported suffering from chronic disease. This study was approved by the Ethics Committee of Binzhou Medical University (Approval number: 2021-337).

### Measures

#### Health literacy

This study employed the All Aspects of Health Literacy Scale (AAHLS) to assess patient health literacy. Specifically designed for primary health care settings, this self-administered scale comprises 14 items categorized into four distinct dimensions: corresponding to skills in using written health information, communicating with health care providers, health information management, and appraisal assertion of individual autonomy with regards to health [35]. Items 1 through 12 were recorded on a 3-point Likert scale, where 1 indicated “rarely” and 3 denoted “usually”. Conversely, Items 13 and 14 were assessed using dichotomous response, options “yes” or “no”. The total score was calculated as the mean value across all items, with higher scores signifying higher levels of patient health literacy. The Cronbach’s α for this scale was 0.819.

#### Patient participation in health care visits

The Patient Perceived Involvement in Care Scale (PICS), was employed to assess patient participation during health care interactions [36]. This 13-item instrument consists of three subscales: doctor facilitation of patient involvement, patient information provision, and patient participation in decision-making. Each item was recorded on a 5-point Likert scale ranging from 1 (“strongly disagree) to 5 (“strongly agree”). The total score was derived from the sum of all item scores. Higher scores indicate greater patient participation in the care process. The Cronbach’s α for this scale was 0.853.

#### Other variables

According to prior research [9, 37–39], additional participant data were collected, including sociodemographic variables (gender, age, marital status, residency, ethnicity, occupational status, education level, and personal monthly income) and health factor (self-rated health status).

### Statistical Analysis

Statistical analyses were performed with IBM SPSS version 24.0 and Mplus version 8.3. First, LPA was employed to identify potential latent classes of patient health literacy. Commonly used model fit indices for LPA include the Akaike information criterion (AIC), Bayesian information criterion (BIC), adjusted Bayesian information criterion (aBIC), Lo-Mendell-Rubin likelihood ratio test (LMR), Bootstrapped likelihood ratio test (BLRT), and Entropy index. Lower values of AIC, BIC, and aBIC suggest a more appropriate model fit. A significant p-value for LMR and BLRT indicates that the k-class model outperforms the (k−1)-class model. Higher Entropy values reflect greater classification accuracy, with an Entropy value ≥ 0.80 corresponding to approximately 90% classification accuracy. While the model fit indices were prioritized, the final number of classes was determined by considering both the practical interpretability of the classification and the sample size within each class. Subsequently, ANOVA was conducted to explore the influence of sociodemographic and health characteristics on the latent profiles of health literacy. Finally, multiple linear regression analysis was performed to assess the impact of health literacy profiles on patient participation.

## Results

### Participant characteristics

Table 1 displays the sociodemographic and health characteristics of the participants. Overall, 45.33% of participants are male, and 54.67% are female. The average age of the participants was 59.26 ± 14.32 years. The majority were married (79.80%), and had a rural household registration (76.40%). A total of 377 participants (41.38%) had a job, and 781 participants (85.73) had an educational level of junior high school or below. Regarding personal monthly income, 73.55% of the participants earned 3000 RMB or less. In addition, 185 participants (20.31%) self-reported their health status as poor.

**Table 1.** Sociodemographic characteristics of the study participants (n=911)

| Characteristic |  | n | % |
| --- | --- | --- | --- |
| Gender | Male | 413 | 45.33 |
|  | Female | 498 | 54.67 |
| Age | (Mean±SD) | 59.26±14.32 | |
|  | <60 years | 453 | 49.73 |
| | ≥60 years | 458 | 50.27 |
| Marital status | Not married | 41 | 4.50 |
|  | Married | 727 | 79.80 |
|  | Divorced or widowed | 143 | 15.70 |
| Residency | Rural | 696 | 76.40 |
|  | Urban | 215 | 23.60 |
| Ethnicity | Han | 903 | 99.12 |
|  | Others | 8 | 0.88 |
| Occupational status | Have no job | 534 | 58.62 |
|  | Have a job | 377 | 41.38 |
| Educational level | Senior high school or below | 781 | 85.73 |
|  | College or above | 130 | 14.27 |
| Personal monthly income | ≤3000 RMB | 670 | 73.55 |
|  | >3000 RMB | 241 | 26.45 |
| Self-rated health | Good | 287 | 31.50 |
|  | Moderate | 439 | 48.19 |
|  | Poor | 185 | 20.31 |

### Latent profile analysis of health literacy

To comprehensively illustrate the distributional differences in health literacy among patients with chronic diseases, this study utilized 14 health literacy items as observed variables and constructed LPA models with one to five potential classes. Table 2 details the fitting results of the LPA models. From Model 1 to Model 5, the AIC, BIC, and aBIC values decreased progressively. Both Model 5 and Model 3 exhibited higher entropy values compared to the other models. However, the LMR value for Model 5 was not statistically significant (*P*>0.05), and the smallest subgroup size in Model 5 accounted for only 5.9% of the sample, which is less than 10%, rendering the accuracy rate of classification not high [40]. After a comprehensive evaluation of the model fit indices, practical significance, interpretability and simplicity of the model, the three-profile model was selected as the optimal model.

**Table 2.** Model fit indices of the latent profile analysis of patient health literacy (n=911)

| Model | AIC | BIC | aBIC | Entropy | LMR (P) | BLRT (P) | Conditional Probability |
| --- | --- | --- | --- | --- | --- | --- | --- |
| 1 | 27241.231 | 27376.038 | 27287.114 |  |  |  |  |
| 2 | 24207.134 | 24414.159 | 24277.597 | 0.917 | <0.001 | <0.001 | 0.637/0.363 |
| 3 | 23172.782 | 23452.025 | 23267.825 | 0.944 | 0.0355 | <0.001 | 0.110/0.569/0.321 |
| 4 | 22296.704 | 22648.165 | 22426.327 | 0.943 | 0.0034 | <0.001 | 0.116/0.186/0.510/<br>0.188 |
| 5 | 21670.679 | 22094.359 | 21814.883 | 0.945 | 0.0903 | <0.001 | 0.059/0.101/0.397/<br>0.151/0.292 |

AIC=Akaike Information Criterion; BIC=Bayesian Information Criterion; aBIC=adjusted BIC; LMR=Lo-Mendell-Rubin likelihood ratio test; BLRT=Bootsrapped likelihood ratio test.

Based on the LPA results, the mean scores of the three profiles across the health literacy items are illustrated in Figure 1. Profile 1 comprised 100 patients (10.98%), with most health literacy items scoring the lowest, thus it was labeled the “Low Health Literacy Group”. Profile 2, which includes 518 patients (56.86%), exhibits moderate scores across the health literacy items and was designated as the “Moderate Health Literacy Group”. Profile 3 consisted of 293 patients (32.16%) who scored the highest on most health literacy items, and was classified as the “High Health Literacy Group”.

**Fig. 1.**
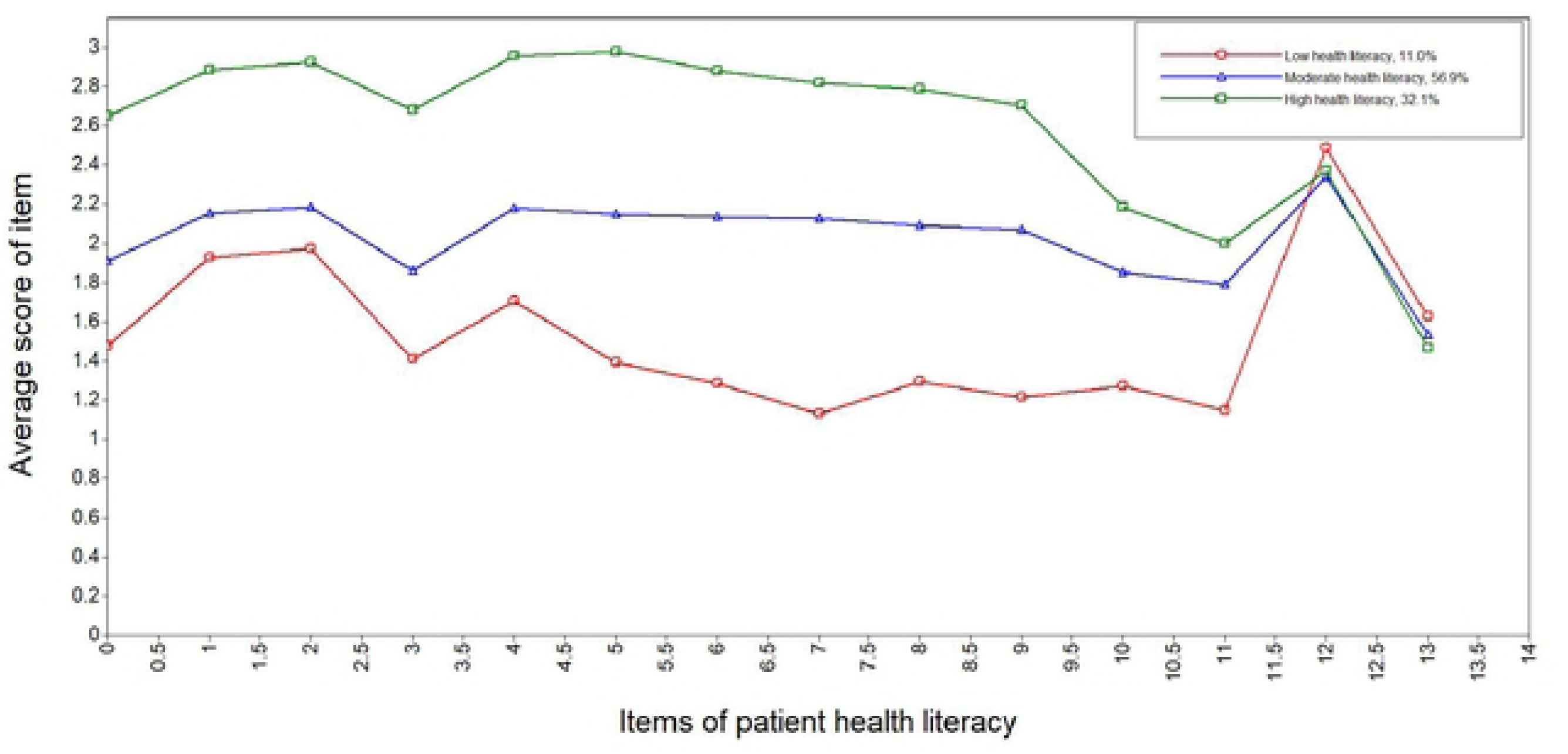
The latent profiles of patient health literacy (n=91I).

### Factors affecting the patterns of health literacy

Table 3 indicates that age, marital status, residency, occupational status, education level, personal monthly income, and health status had statistically significant effects on the latent profiles of patient health literacy (*P*<0.05).

**Table 3.** Sociodemographic characteristics associated with the three latent profiles of patient health literacy.

| Characteristic | Low health literacy | Moderate health literacy | High health literacy | $\chi^2$ | *P* value |
| --- | --- | --- | --- | --- | --- |
| Gender |  |  |  |  |  |
| Male | 42(10.17) | 223(54.00) | 148(35.84) | 4.709 | 0.095 |
| Female | 58(11.65) | 295(59.24) | 145(29.12) |  |  |
| Age |  |  |  |  |  |
| <60 years | 39(8.61) | 301(66.45) | 113(24.94) | 33.756 | <0.001 |
| ≥60 years | 61(13.32) | 217(47.38) | 180(39.30) |  |  |
| Marital status |  |  |  |  |  |
| Not married | 6(14.63) | 26(63.41) | 9(21.95) | 20.247 | <0.001 |
| Married | 74(10.18) | 434(59.70) | 219(30.12) |  |  |
| Divorced or widowed | 20(13.99) | 58(40.56) | 65(45.45) |  |  |
| Residency |  |  |  |  |  |
| Urban | 11(5.12) | 148(68.84) | 56(26.05) | 19.178 | <0.001 |
| Rural | 89(12.79) | 370(53.16) | 237(34.05) |  |  |
| Ethnicity |  |  |  |  |  |
| Han | 100(11.07) | 513(56.81) | 290(32.12) | 1.003 | 0.606 |
| Others | 0(0.00) | 5(62.50) | 3(37.50) |  |  |
| Occupational status |  |  |  |  |  |
| Have no job | 79(14.79) | 293(54.87) | 162(30.34) | 19.365 | <0.001 |
| Have a job | 21(5.57) | 225(59.68) | 131(34.75) |  |  |
| Educational level |  |  |  |  |  |
| Senior high school or below | 93(11.91) | 435(55.70) | 253(32.39) | 6.521 | 0.038 |
| College or above | 7(5.38) | 83(63.85) | 40(30.77) |  |  |
| Personal monthly income |  |  |  |  |  |
| ≤3000 RMB | 81(12.09) | 357(53.28) | 232(34.63) | 13.338 | 0.001 |
| >3000 RMB | 19(7.88) | 161(66.80) | 61(25.31) |  |  |
| Self-perceived health status |  |  |  |  |  |
| Good | 36(12.54) | 165(57.49) | 86(29.97) | 22.235 | <0.001 |
| Fair | 38(8.66) | 274(62.41) | 127(28.93) |  |  |
| Poor | 26(14.05) | 79(42.70) | 80(43.24) |  |  |
| Total | 100(10.98) | 518(56.86) | 293(32.16) | - | - |

These significant variables were subsequently included in the multinomial logistic regression model to determine the predictors associated with each health literacy profile. Compared with patients in the high health literacy group, those who have no job were more likely to be categorized into the low health literacy group (OR = 2.870, 95% CI = 1.562-5.274). Meanwhile, patients under 60 years old (OR = 1.925, 95% CI = 1.388-2.672), urban residents (OR = 1.725, 95% CI = 1.134-2.624), individuals that have no job (OR = 1.654, 95% CI = 1.157-2.365), and those who self-reported good health status (OR = 1.585, 95% CI = 1.031-2.437) were more inclined to fall into the moderate health literacy group. In addition, patients with a monthly income of 3,000 yuan or less were less likely to be classified into the moderate health literacy group (OR = 0.566, 95% CI = 0.391-0.821). Specific details are presented in Table 4.

**Table 4.** Multinomial logistic regression for patient health literacy profiles.

| Characteristics | Low health literacy |  |  | Moderate health literacy |  |  |
| --- | --- | --- | --- | --- | --- | --- |
|  | *B* | *OR* | 95%*CI* | *B* | *OR* | 95%*CI* |
| Age (Reference: $\geq 60$ years) | | | | | | |
| <60 years | 0.190 | 1.210 | (0.719,2.034) | 0.655 | 1.925 | (1.388,2.672) |
| Marital status (Reference: Divorced or widowed) |  |  |  |  |  |  |
| Not married | 1.026 | 2.789 | (0.711,10.936) | 0.525 | 1.690 | (0.659,4.330) |
| Married | 0.247 | 1.280 | (0.695,2.356) | 0.481 | 1.618 | (1.056,2.481) |
| Residency (Reference: Rural) |  |  |  |  |  |  |
| Urban | -0.054 | 0.947 | (0.429,2.089) | 0.545 | 1.725 | (1.134,2.624) |
| Occupational status (Reference: Have a job) |  |  |  |  |  |  |
| Have no job | 1.054 | 2.870 | (1.562,5.274) | 0.503 | 1.654 | (1.157,2.365) |
| Educational level (Reference: College or above) |  |  |  |  |  |  |
| Senior high school or below | 0.689 | 1.991 | (0.674,5.883) | 0.358 | 1.430 | (0.859,2.382) |
| Personal monthly income (Reference: >3000 RMB) |  |  |  |  |  |  |
| $\leq 3000$ RMB | -0.156 | 0.855 | (0.462,1.584) | -0.568 | 0.566 | (0.391,0.821) |
| Self-rated health (Reference: Poor) |  |  |  |  |  |  |
| Good | 0.235 | 1.264 | (0.684,2.337) | 0.461 | 1.585 | (1.031,2.437) |
| Fair | -0.084 | 0.919 | (0.505,1.675) | 0.673 | 1.960 | (1.313,2.925) |

Ref: High health literacy

### Relationship between health literacy profiles and patient participation

Table 5 reflects that the overall average score of patient participation was 10.50±3.01. The results of the ANOVA revealed that patients with low health literacy had significantly lower scores in doctor facilitation (F=34.093), patient information(F=72.100), patient decision-making(F=20.474), and overall patient participation (F=57.255) compared to those with moderate or high health literacy(P<0.001).

**Table 5.** Association between health literacy profiles and patient participation.

| Subgroups of patient health literacy | Doctor facilitation | Patient information | Patient decision-making | Patient participation |
| --- | --- | --- | --- | --- |
| Low health literacy | 3.49±1.61 | 2.41±1.54 | 1.93±1.55 | 7.83±3.98 |
| Moderate health literacy | 4.37±1.12 | 3.40±1.08 | 2.76±1.43 | 10.53±2.92 |
| High health literacy | 4.58±0.99 | 3.80±0.52 | 2.96±1.27 | 11.34±2.13 |
| Total | 4.34±1.19 | 3.42±1.08 | 2.73±1.43 | 10.50±3.01 |
| *F* | 34.093 | 72.100 | 20.474 | 57.255 |
| *P* | <0.001 | <0.001 | <0.001 | <0.001 |

The simple linear regression model was applied to primarily examine potential socioeconomic and health factors influencing patient participation. Significant relationships were found as follows: self-rated health and doctor facilitation; marital status, occupational status, self-rated health and patient information; marital status, occupational status, self-rated health and patient decision-making; marital status, occupational status, self-rated health and overall patient participation. Consequently, sociodemographic and health charcateristics showed significance were entered into multiple linear regression models as covariates to adjust for confounding factors.

Table 6 shows the results of the multiple linear regression analysis examining the associations between health literacy profiles and patient participation, as well as its individual domains, after adjusting for related sociodemographic and health characteristics. The results indicated that patients with high health literacy exhibit significantly higher levels of doctor facilitation (B = 1.103, p < 0.001), patient information (B = 1.391, p < 0.001), patient decision-making (B = 1.042, p < 0.001), and overall patient participation (B = 3.544, p < 0.001) compared to those with low health literacy.

**Table 6.**
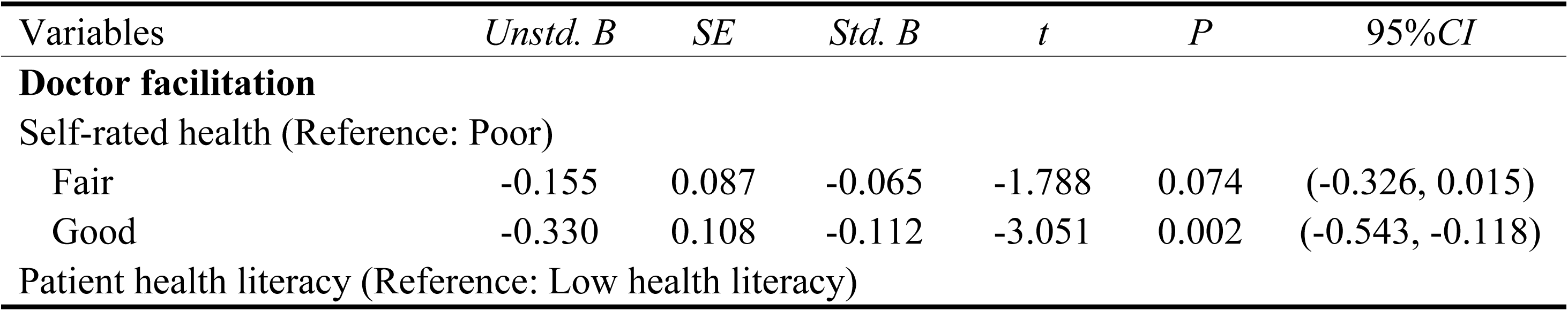
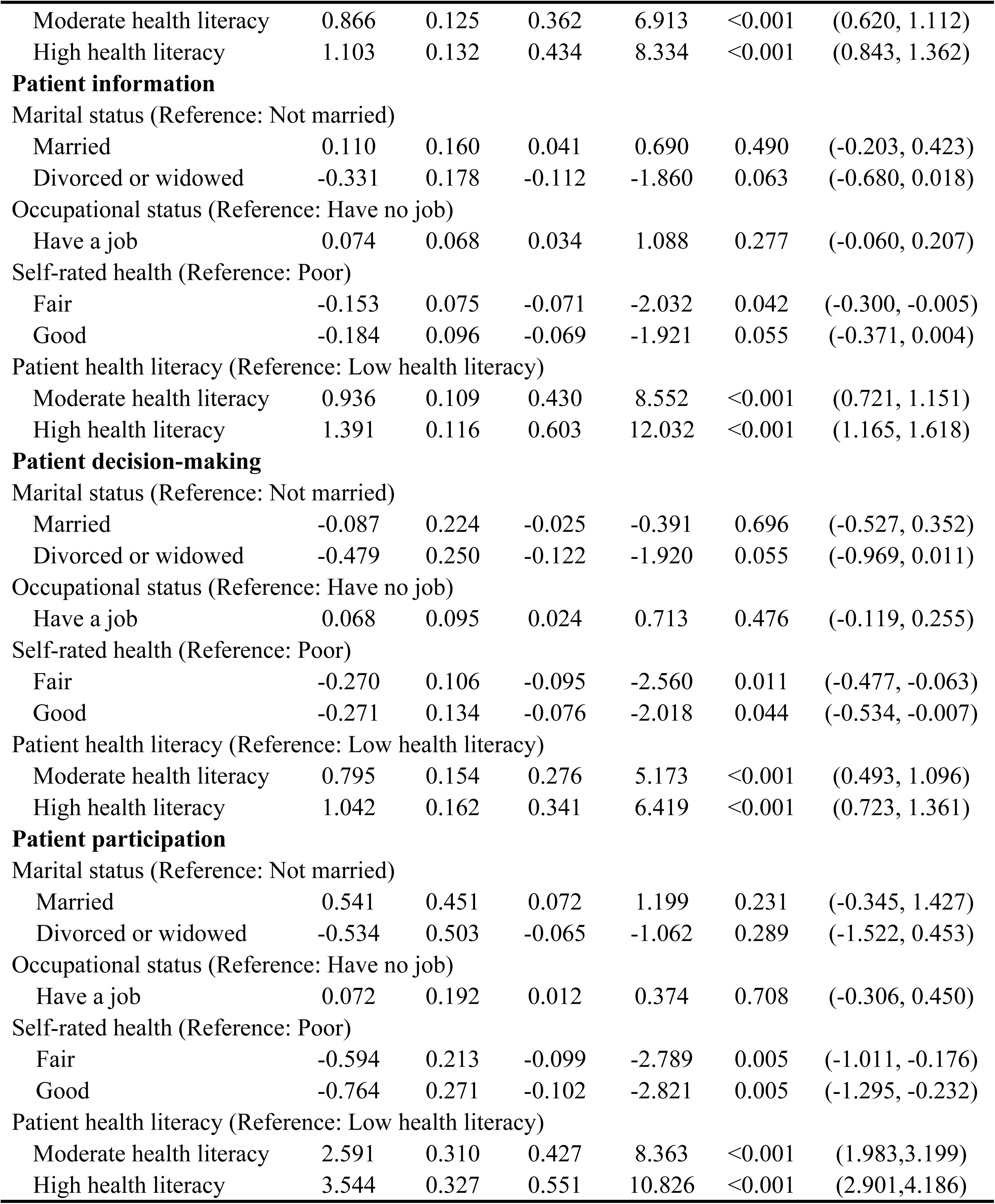
Multiple linear regression model for patient participation.

## Discussion

In the present study, we investigated the profiles of patient health literacy and their association with patient participation in primary health care through a cross-sectional survey of 911 patients with chronic diseases in Shandong Province. Our findings revealed that health literacy among chronic disease patients exhibited heterogeneity, and displayed distinct patterns. Three different categories were identified: low health literacy group, moderate health literacy group, and high health literacy group. Furthermore, age, marital status, residency, occupational status, education level, personal monthly income, and health status were significant determinants of these latent subgroups. The findings also indicated that patients with high or moderate health literacy were more likely to exhibit higher participation in primary health care compared to those with low health literacy. To our knowledge, this is the first study dedicated to employ a rigorous scientific method to discern the patterns of patient health literacy, and examine how these categories influence participation in primary health care among patients with chronic diseases. These findings contribute to the research on patient participation, offering new evidence that higher levels of health literacy have a positive impact on patient participation in primary health care. Moreover, the findings may serve as a guide for health care managers and policymakers to improve health literacy among patients with chronic diseases, and promote their participation in primary health care.

An analysis of patient health literacy profiles revealed three distinct classifications: low health literacy, moderate health literacy, and high health literacy, emphasizing the heterogeneity of health literacy among patients with chronic diseases. Notably, our findings indicate that 67.84% of the participants fell into the moderate or low health literacy group, which is in line with findings from previous studies showing that health literacy among patients with chronic diseases tends to be low [41–43]. This is particularly concerning given the nature of chronic diseases, which require long-term management, adherence to treatment plans, and the sustained adoption of healthy lifestyle practices. Effective self-management, in turn, demands an understanding of complex health information and the ability to apply them in daily life [44]. Health literacy, which encompasses the knowledge, motivation, and skills necessary to obtain, comprehend, evaluate, and utilize health information, is essential for making informed decisions aimed at maintaining or enhancing one’s health and quality of life [26, 43]. Given its crucial role in chronic disease prevention and control [42, 44, 45], health care managers and policymakers should prioritize initiatives aimed at improving health literacy among patients with chronic diseases.

In terms of the determinants of patient health literacy profiles, our analysis identified several significant factors, including age, marital status, residency, occupational status, education level, personal monthly income, and health status. These findings resonate with other studies indicating that patients aged 60 years or older, had no job or with low-income are more likely to have limited health literacy [24, 42, 46]. Regarding education, results showed that patients with senior high school or below exhibited lower health literacy than those with college or above. One possible explanation for this is that patients with higher education levels generally have better access to health information, enhanced critical thinking and communication skills, greater health awareness, and exposure to formal health education [41]. Furthermore, our findings revealed that rural patients were more likely to have lower health literacy than urban patients, which is consistent with previous research [42]. One reason for this disparity could be the limited availability of educational and health care resources in rural areas, coupled with restricted access to health-related information. These challenges may contribute to the observed gap in health literacy observed between rural and urban populations [47]. In addition to socioeconomic and demographic factors, health status also emerged as a significant determinant of patient health literacy, which is in line with previous findings [41, 48]. These results highlight the need for tailored interventions to improve health literacy among patients with chronic diseases.

Regarding the participation among patients with chronic disease, the average score was 10.50±3.01 (out of a total score of 13), which is notably higher than the previously reported scores of 6.71±3.35 from China’s northeastern province [11]. This discrepancy may be related to various geographic, economic and policy-related factors that influence the capability of primary health care facilities to deliver services. In addition, differences in survey sample selection may have significantly contributed to the observed variations. Given the complexity of chronic disease, active patient involvement is essential for effective disease management. Participation in primary health care delivery plays a crucial role in early detection, treatment adherence, and improved health outcomes [11, 25]. Furthermore, promoting patient participation in medical encounters fosters trust in physicians [49], enhances patient satisfaction, and increases their willingness to seek and use health services [13]. Therefore, health care managers and policymakers should place significant emphasis on patient participation in chronic disease management, and establish supportive policies and guidelines to enhance patient participation during the process of primary health care delivery. This could include the widespread promotion of shared decision-making, training physicians to develop patient-centered communication skills, and raising patients’ awareness about the benefit of active participation in health care interactions.

With respect to the relationship between health literacy profiles and patient participation, our results showed that patients with higher health literacy exhibited significantly greater participation in primary health care visits compared to those with lower health literacy. While health literacy has been widely recognized as a key determinant of patient participation in decision-making, its specific impact on participation in primary health care among patients with chronic diseases remains underexplored. Our findings provide valuable empirical evidence supporting the role of health literacy in improving patient participation in primary health care. This study contributes to the growing body of literature on patient participation in primary health care for chronic disease management, and underscores the importance of health literacy as a facilitator of active patient participation. Therefore, health care managers and policymakers should take proactive steps to promote health literacy, particularly among individuals with chronic diseases, and ensure equitable access to health literacy initiatives. Potential interventions include developing tailored health education programs that address the specific needs of patients with chronic diseases, and leveraging digital health applications to provide patients the opportunity to acquire reliable health information.

### Limitations and future research

To the best of our knowledge, this study represents the first attempt to investigate the relationship between health literacy and participation in primary health care among patients with chronic diseases, in China, which provide valuable insights for future research in this area. Simultaneously, several limitations in this study ought to be noted. First, the findings of this cross-sectional study were correlational in nature, as such no causal conclusions can be drawn. Future research should investigate this topic adopting experimental methods or longitudinal study designs. Second, owing to the constraints related to time, funding, and human resource, our data collection was limited to a single province in China. As a result, the generalizability of our findings may be limited. Therefore, future studies should extend the study area to cover other geographic regions to improve the generalizability of the research conclusions. Third, all data collected were self-reported, which may introduce recall bias. Although the interviewers were trained in appropriate techniques to assist participants provide accurate responses.

## Conclusion

This study explored the relationship between health literacy and patient participation in primary health care among individuals with chronic diseases, contributing to the growing body of literature on primary health care. These findings indicate that health literacy plays a pivotal role in shaping patient participation in health care interactions. In addition, these findings offer valuable implications for recognizing health literacy as a key factor in promoting patient participation. Therefore, health care managers and policymakers should prioritize efforts to improve health literacy among patients with chronic diseases. This includes promoting equitable access to health literacy programs, and designing tailored interventions for optimizing patient health literacy.

## Data Availability

Data are available upon reasonable request.

## Author contributions

Conceptualization: Qi Xu, Li Zhang.

Data curation: Qi Xu, Baokai Wang.

Formal analysis: Baokai Wang, Li Zhang.

Investigation: Baokai Wang, Li Zhang.

Visualization: Qi Xu, Li Zhang.

Writing – original draft: Qi Xu, Li Zhang.

Writing – review & editing: Li Zhang.

## Acknowledgements

We would like to thank all participants in the study and the organizations that supported the project.

